# Prevalence and factors associated with depressive symptoms among adults with glaucoma at a tertiary hospital in Tanzania: A cross-sectional study

**DOI:** 10.64898/2026.02.26.26347156

**Authors:** Jawaher Soud Rashid, Samuel Chacha, Florian Emmanuel Ghaimo, Ester Steven Mzilangwe, Zahra Morawej, Celina Mhina, Said Bakari Kuganda

## Abstract

**Background:** Glaucoma is identified as one of the leading causes of blindness worldwide. Its chronic nature and the potential for irreversible vision loss contribute to significant distress among affected individuals. Around 25% of individuals with glaucoma are estimated to experience depression, negatively impacting their quality of life and treatment adherence. However, data on the prevalence of depression among people with glaucoma in Tanzania is limited. This study aimed to determine the prevalence and factors associated with depressive symptoms among adults with glaucoma at Muhimbili National Hospital.

**Materials and methods:** A cross-sectional study was conducted involving 297 adults with glaucoma, who were recruited consecutively from the ophthalmology clinic at Muhimbili National Hospital between July and November 2024. Data on biopsychosocial factors were collected using interviewer-administered questionnaires and medical records. Patient Health Questionnaire-9 and Oslo Social Support Scale assessed depressive symptoms and social support, respectively. Data were analyzed using STATA version 16. Logistic regression analyses identified factors associated with probable depression, with statistical significance set at p-value<0.05.

**Results:** The mean age of participants was 63.6 years (SD±12.8), with 159 (53.5%) being female. Prevalence of probable depression was 11.1%, with 8.7% moderate, 2.4% moderately severe, and none reporting severe depressive symptoms. Having moderate social support (AOR 0.14; CI: 0.04–0.47; P=0.001) and strong social support (AOR 0.08; CI: 0.03–0.25; P<0.000) were significantly associated with lower odds of probable depression.

**Conclusion:** Approximately 1 in 10 individuals with glaucoma experience depression. Having good social support was identified as a protective factor against depression in people with glaucoma. These findings underscore the need for a multidisciplinary approach integrating psychosocial services into ophthalmology clinics.

## Introduction

Glaucoma is a major cause of irreversible blindness worldwide (1), ranking second among all causes of blindness(2) following cataract. Estimates suggest that the number of individuals affected by glaucoma will rise to 111.8 million by 2040 (1). Glaucoma is an irreversible optic neuropathy that causes structural changes at the optic nerve head, which progressively cause severe visual field loss. In 2013, Africa had the second highest number of individuals with glaucoma (1), with the most recent prevalence of glaucoma reported at 5.59% (3). In rural central Tanzania, the estimated prevalence of glaucoma is 4.16% (4). The chronic and progressive nature of glaucoma leads to visual disability, which interferes with daily activities (5), and may in turn contribute to the development of depression (6).

Chronic medical conditions like glaucoma heighten the risk of depression and, in turn, impact medical treatment outcomes (7). Individuals with glaucoma often have co-morbid psychiatric disorders, most often anxiety and depression (8,9). Studies from across the globe show varying prevalence of depressive symptoms among persons with glaucoma, ranging between 10 - 57%, including Japan, Turkey, USA, China, USA, Brazil, Germany, and Nigeria (10–15). For example, a population-based cohort study in Germany among 14,657 persons aged 35 to 74 years, with and without glaucoma, using PHQ-9 found a lower prevalence of depression (6.6%), with 24.3% having minimal, 4.9% mild, 1.7% moderate-severe, and 0.0% severe depressive symptoms (16). In Turkey, a case-control study of 121 adults found a notably higher prevalence of 57% (11). A study in Australia using the Geriatric Depression Scale (GDS-15) among 68 participants aged above 60 years found that the prevalence of depression increases with increased severity of glaucoma: 11.4% in those with mild, 20.9% with moderate, and 32.1% with severe glaucoma (17). While in Africa, few studies have explored depression among individuals with glaucoma. Most available studies are from Nigeria, like the case-control study using Hospital Anxiety and Depression Scale (HADS), Ubochi et al. found a prevalence of 21.7% (18). A more recent comparative study by Okudo et al. revealed that among adults with a diagnosis of glaucoma for at least 6 months, the prevalence of depression was reported at 24.4%, with 12.7% having mild, 5.6% moderate, and 6.1% severe depressive symptoms (19).

Individuals with glaucoma experiencing depressive symptoms may exhibit poor adherence to clinical recommendations (12), elevating their risk of vision loss and therefore affecting the person’s ocular and overall health. If left untreated, depressive symptoms may lead to the subsequent development of major depression, impairing daily functioning, increasing dependence on others, and diminishing quality of life (QoL) (20), and increasing the healthcare system’s and the community’s economic burden (15). Individuals with glaucoma in Tanzania already face significant challenges with treatment adherence (21), and the presence of depressive symptoms may further exacerbate this problem, elevating their risk of vision loss and negatively impacting both ocular and overall health.

Several potential risk factors for depression among persons with glaucoma were identified, including older age (6,15,17,22), female sex (6,13,14,23), being separated or widowed (15), living alone (24), poor social support (25), increased glaucoma severity (6,13,17,22,23,26), and a longer duration of glaucoma (11,15). The psychosocial needs of people with glaucoma are frequently overlooked, leading to potential underdiagnosis and under-treatment of psychiatric comorbidities. Societal stigma surrounding mental health conditions and the limited awareness among both patients and ophthalmologists may further contribute to the under-treatment, hence hindering access to proper psychological care (23).

The chronic nature of glaucoma and its association with an increased risk of depression raise its public health significance and highlight the need to explore the burden of depression and associated factors within our setting. A multidisciplinary approach is required to identify, assess, and refer patients to specialized mental health care providers for further management and improvement of their quality of life (QoL).

Despite the vast amount of research done on the magnitude and factors associated with depression among persons with glaucoma elsewhere in the world, there is no retrievable information in the Tanzania context to the best of our knowledge. Therefore, this study aims to examine the prevalence and factors associated with depressive symptoms among people with glaucoma.

## Methods

### Study Design and setting

This is a facility-based, analytical, cross-sectional design that employed a quantitative method to determine the prevalence of depressive symptoms and associated factors among adults with glaucoma attending the ophthalmology clinic at Muhimbili National Hospital. The Muhimbili National Hospital is a tertiary referral hospital located in Dar-es-salaam region of Tanzania, which is the commercial city of Tanzania(27), it receives patients from different regions in the country. It is a tertiary hospital offering specialized medical services, both inpatient and outpatient services. It has a 1,500-bed facility and attends around 1,000 to 1,200 outpatients per week. The ophthalmology department has glaucoma clinics that provide access to people with glaucoma inpatient and follow-up clinics that run weekly on Tuesdays. The clinic receives around 160 – 240 patients per month and is operated by a consultant ophthalmologist, nurses, and resident doctors. The services offered at the glaucoma clinic include medication, laser therapy, trabeculoplasty, glaucoma implant valves, cyclophotocoagulation etc.

### Selection criteria

The study included all adults with a confirmed diagnosis of glaucoma in the past six months (19) aged 18 years and above, attending the ophthalmology clinic at MNH during the study period, and consented to take part in the research. Participants with critical illness who are incapable of communicating were excluded from the study.

### Sample size estimation

The minimum sample size estimated was 312 using Cochran’s formula (28). Adjustment for 10% non-response rate was considered. The assumption of prevalence used was 24.4%, obtained from a study in Nigeria on the prevalence of depression among persons with glaucoma (19).

### Sampling and recruitment

We used a consecutive non-probability sampling method, where all eligible adults attending the glaucoma clinic present during the study period were selected consecutively, approached, and invited to participate until the desired sample size was reached. This sampling technique was chosen due to the limited time available to conduct the study and the limited number of patients at the study site, as these are the same patients who return for follow-up. Of the 312 participants enrolled, 297 completed the study, resulting in a response rate of 95.2%. Ten participants could not continue with the interviews due to time constraints. Additionally, data from five participants were incomplete because the glaucoma severity could not be reported due to hazy media that resulted in difficulty in visualizing of fundus during the eye examination to measure the cup-to-disk ratio. Figure 1 shows the flow of participants through the study.

**Fig. 1:**
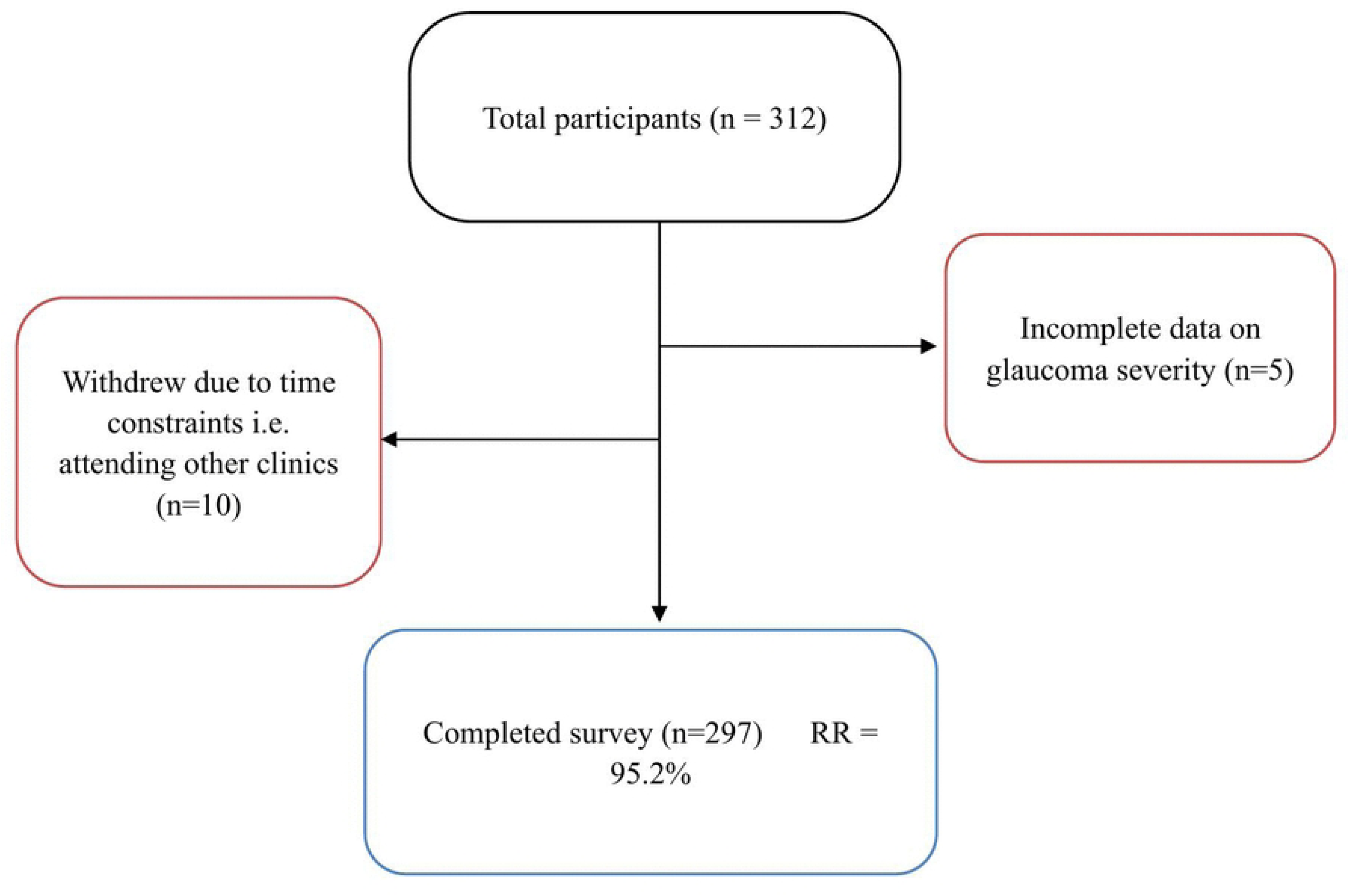
Flow chart of participants through the study.

### Data collection instruments and procedures

Data collection was through a face-to-face, interviewer-administered structured questionnaire, which was translated from English, the original version, to Swahili. Swahili was chosen because it’s the main language used for communication and is widely understood by most people in Tanzania. The questionnaires were piloted and administered by the principal investigator and two research assistants with medical credentials. Prior data collection, two days orientation training was conducted for the research assistants to standardize the administration of questionnaires. The questionnaire was programmed into the software program Kobo Toolbox, where they were administered, and responses were recorded in the platform using a tablet device. Interviews were conducted in a private setting to ensure confidentiality from July 2024 to November 2024. Regular supervision was performed to ensure data quality and completeness.

### Study measures

Key variables in the questionnaire were based on the existing literature, which covered the sociodemographic, clinical characteristics, participants’ medical record (severity of glaucoma and eye co-morbidities), assessment of depression using Patient Health Questionnaire −9, and social support using Oslo Social Support Scale.

#### Socio-demographic information and clinical characteristics

The key variables of socio-demographic and clinical characteristics were developed by the researcher based on factors associated with depression and depressive symptoms from previous literature. These factors included age, sex, level of education, marital status, employment status, who they live with (alone, spouse or other family members), self-reported history of comorbid medical conditions (hypertension, diabetes mellitus, stroke and cancer) (16,29), use of substances in the past 12 months such as alcohol consumption and cigarette smoking/tobacco use (24), duration of glaucoma (less than 5 years ago or 5 years), the number of a person’s current anti-glaucoma medications (less than three or more medications) (15,17,24) and history of glaucoma surgery in the past six months. Eye comorbidities of participants were obtained from their medical records documents by the attending doctors. The level of glaucoma severity was obtained from the recorded measurements done by an ophthalmologist using a slit lamp and handheld retina lens. Severity was graded based on the cup-to-disc ratio, with a ratio of less than 0.5 classified as normal, 0.5 to 0.6 as mild, 0.7 to 0.8 as moderate, and greater than 0.9 as advanced glaucoma (30).

#### Depressive symptoms

Depressive symptoms were assessed using the Patient Health Questionnaire (PHQ-9). The PHQ-9 was developed by Kroenke et al and it is composed of 9 items from the DSM V criteria of depression, for each item, it had scores ranging from “0” to “3” (nearly every day) (0 indicating not at all, 1 – a few days, 2 – more than half the days and 3 - nearly every day) (31). Participants reported how often they experienced the symptom in the item over the previous two weeks. Total scores range from 0 to 27, whereby the scores are categorized into non or minimal, mild, moderate, moderate-severe, and severe for scores ranging from 0 – 4, 5 – 9, 10 – 14, 15 – 19, and 20 – 27, respectively (31). A validity study among primary care outpatients in Tanzania found that the PHQ-9 had a sensitivity of 78% and a specificity of 87% for major depression, with reasonable reliability (Cronbach α of 0.83), and a cut-off point of a score greater than or equal to 9 was considered optimal in this population (32). Individuals who scored 9 or more were categorized as having depression.

#### Social support

We used Oslo Social Support Scale (OSSS-3) to measure social support among participants. Some of the constructs in the scale are related to mental health issues, particularly depressive symptoms (33). This tool was also utilized in other studies (9) to measure the level of social support among persons with glaucoma and common mental health disorders. The tool has three items: the presence of confidants a person can turn to, the sense that other people are concerned about your life, and the relationship with neighbors to access help. Each item is assigned scores, with the total sum of all scores equaling 14. Scores for each category include 3 – 8, 9 – 11, and 12 – 14, indicating “poor”, “moderate”, and “strong” social support, respectively. The OSSS-3 was shown to have an internal consistency of Cronbach’s α of 0.64 (9) in a population with glaucoma. Still, it has demonstrated good reliability (Cronbach α 0.88) in other populations (8). Although it has not been validated in the Tanzanian cultural context, it has been used in Tanzanian settings (34).

### Data Processing and Analyses

Data were exported from Kobo Toolbox to Microsoft Excel for cleaning. The cleaned data were then imported into STATA version 16 (35) in codes for processing and analysis. Descriptive statistics were summarized using frequencies and percentages for categorical variables, and mean and standard deviation for continuous variables. The results were summarized and presented in tables and figures. Bivariable were performed to assess association between independent variables and depression, where Pearson Chi-Square test was used for categorical variables. For variables with expected counts less than 5, Fisher’s exact test was applied and these variables were subsequently included in the multivariable analysis to control for potential confounding. Variables with a P-value < 0.2 in bivariable analysis were further selected for multivariable regression analysis. The odds ratio with 95% confidence intervals (CI) was reported to show the association between independent variables of interest and probable depression. Factors were deemed statistically significant if their P-value was < 0.05. Correlation and intercorrelation analyses were performed to assess for multicollinearity among independent variables.

### Ethical Consideration

Approval for the study was acquired from the Institutional Review Board of Muhimbili University of Health and Allied Sciences (MUHAS-REC-06-2024-2312). Permission to carry out the study was requested from the MNH Research and Training Unit, and approval to conduct the research was sought from the Head of the Department of Ophthalmology at MNH. Consent forms were provided to all participants (see S1 File), and those found to have depression were linked and directed to mental health services.

## Results

### Sociodemographic and clinical characteristics of study participants

The mean age of the study participants was 63.6 years (SD ± 12.8), with nearly three-quarters (72.4%) aged above 60 years. Over half of the participants (53.5%) were females, and nearly two-thirds (62.3%) were married. Almost one-third (32.0%) were divorced or separated. In addition, over three-quarters (88.9%) of the participants lived with family members, and nearly two-thirds (63.6%) reported having strong social support. Table 1 below, summarizes the socio-demographic characteristics of the study participants. Two-thirds (66.0%) of the participants were diagnosed with glaucoma within the past 5 years. Nearly half (45.5%) of the participants had chronic illnesses, with a majority having hypertension (53.3%). Individuals who reported to have used substances within 12 months were 8.7%. Most participants (82.8%) were using fewer than three antiglaucoma medications. Furthermore, 14.8% had undergone eye surgery in the past 6 months, and almost half (48.2%) had advanced glaucoma severity. Table 1 demonstrates the clinical characteristics of the study participants.

**Table 1.** Sociodemographic and clinical characteristics of participants 2024 (n=297)

| Characteristics | Frequency, n (%) |
| --- | --- |
| **Sex** |  |
| Male | 138 (46.5) |
| Female | 159 (53.5) |
| **Age, years: Mean (SD ±)** | 63.6 (±12.8) |
| **Age group, years** |  |
| 18–35 | 11 (3.7) |
| 36–59 | 71 (23.9) |
| ≥ 60 | 215 (72.4) |
| **Marital status** |  |
| Single | 17 (5.7) |
| Married | 185 (62.3) |
| Divorced/Separate | 95 (32.0) |
| **Education level** |  |
| No formal/Primary education | 142 (47.81) |
| Secondary education | 95 (32.0) |
| College/university education | 60 (20.2) |
| **Employment status** |  |
| Unemployed | 190 (64.0) |
| Employed | 107 (36.0) |
| **Living arrangement** |  |
| Alone | 7 (2.4) |
| Spouse | 26 (8.7) |
| Other family members | 264 (88.9) |
| **Social support** |  |
| Poor | 25 (8.4) |
| Moderate | 83 (28.0) |
| Strong | 189 (63.6) |
| **Duration of Glaucoma (years)** |  |
| <5 | 196 (66.0) |
| ≥5 | 101 (34.0) |
| **Comorbid medical illnesses** |  |
| No | 162 (54.6) |
| Yes | 135 (45.5) |
| **History of eye surgery past six months** |  |
| No | 253 (85.2) |
| Yes | 44 (14.8) |
| **Severity of glaucoma** |  |
| Normal | 58 (19.5) |
| Mild | 47 (15.8) |
| Moderate | 49 (16.5) |
| Advanced | 143 (48.2) |
| **Eye comorbidities** |  |
| None | 256 (86.2) |
| Cataract | 26 (8.8) |
| Neovascular glaucoma | 13 (4.4) |
| Dense cataract and neovascular glaucoma | 1 (0.3) |
| Proliferative diabetic retinopathy | 1 (0.3) |

Key: SD = Standard deviation

### Prevalence and severity of depressive symptoms among adults with glaucoma

The prevalence of clinically significant depressive symptoms (PHQ-9 ≥ 9) among persons with glaucoma at MNH was 11.1%. None of the participants reported severe depressive symptoms. Only 2.4% had moderately severe depressive symptoms, 8.7% had moderate depressive symptoms, 11.8% had mild depressive symptoms, and 77.1% showed no/minimal depressive symptoms, as illustrated in Figure 2.

**Fig. 2:**
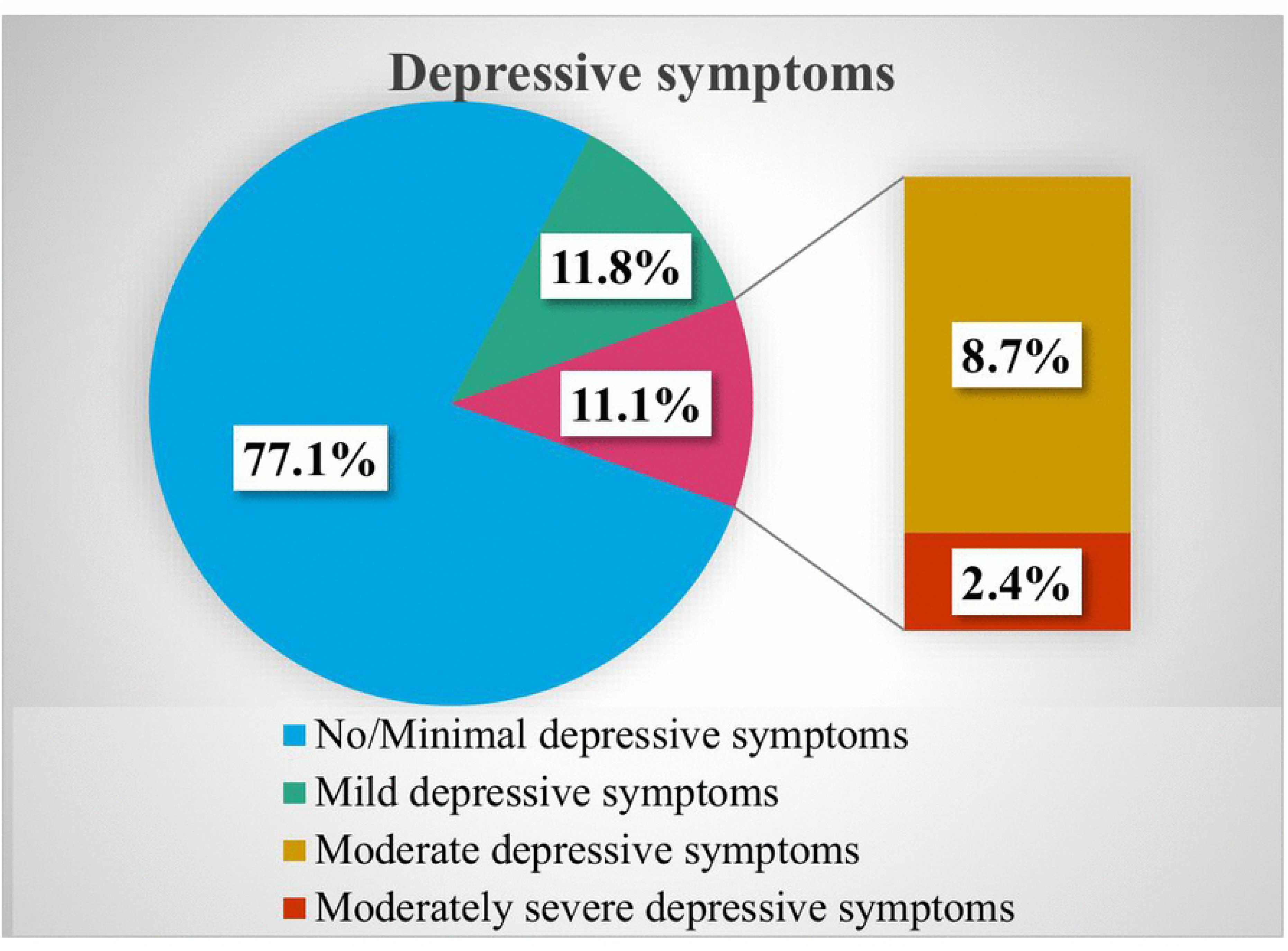
Prevalence of depressive symptoms among adults with glaucoma in a tertiary hospital, Tanzania, 2024.

### Factors associated with depression among adults with glaucoma

The factors found to have a statistically significant association in the bivariable analysis were having a college/university education, living with family members, and having moderate to strong social support.

Participants having a college/university level education showed 64% lower odds (COR=0.36; CI 95% CI: 0.10-1.28; P=0.115) of having probable depression compared to those with no formal/primary level education. The participants who were living with family members had 73% lower odds (COR=0.27; 95% CI: 0.05–1.48; P<0.132) of having probable depression compared to living alone. While those with moderate social support had 85% lower odds (COR=0.15; 95% CI: 0.05–0.44; P<0.001), and those with strong social support had 91% lower odds (COR=0.09; 95%CI: 0.04–0.25; P<0.001) of screening positive for probable depression compared to those with poor social support.

However, in multivariable analysis, having a college/university level education (AOR=0.32; 95% CI: 0.075-1.39; P=0.131) and living with other family members (AOR=0.09; 95%CI: 0.01-1.14; P=0.063) showed no significant association with probable depression. Social support was the only factor found to have statistically significant association with probable depression. Participants having moderate social support had 86% lower odds (AOR=0.14; 95% CI: 0.04–0.47; P< 0.001), and the participants with strong social support had 92% lower odds (AOR=0.08; 95% CI: 0.03–0.25; P<0.000) of having probable depression. All other independent factors were found not to be statistically significant.

These results showed no significant correlations among the predictor variables and sociodemographic factors (e.g., between mental illness and social support, or living with someone and social support), suggesting that multicollinearity was not a concern in the regression analysis. Table 2 shows the bivariable and multivariable analysis.

**Table 2.**
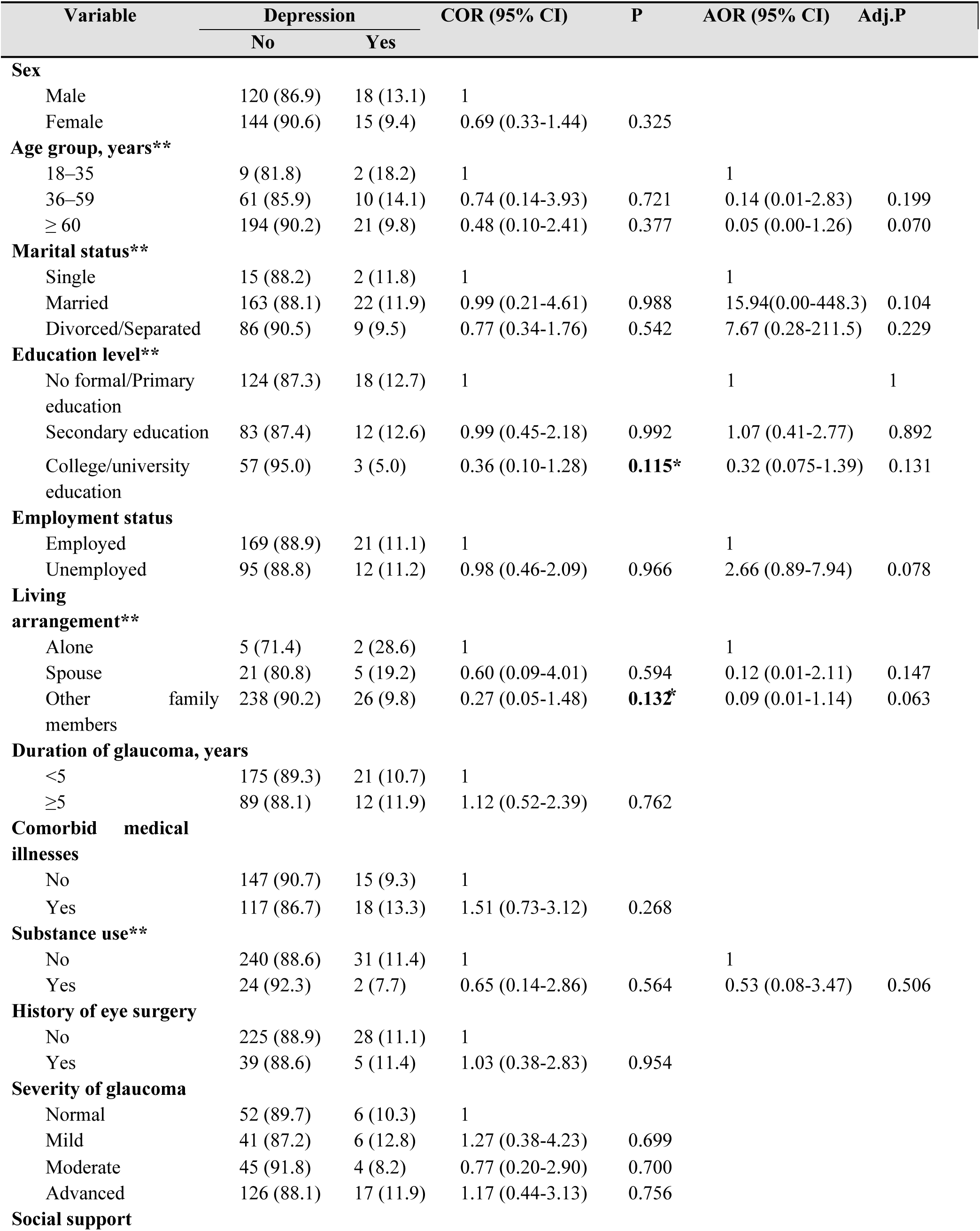
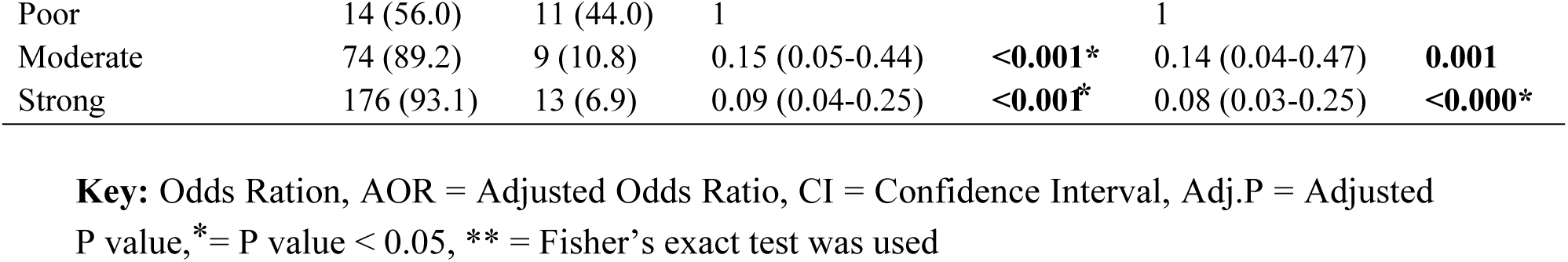
Logistic regression models to determine factors associated with ession among adults with glaucoma.

## Discussion

In our study, the prevalence of depressive symptoms among individuals with glaucoma is 77.1% for no/minimal depressive symptoms, 11.8% mild, 8.7% moderate, 2.4% for moderately severe symptoms, and none for severe depressive symptoms. The overall rate of probable depression at a cut-off point of ≥ 9 was 11.1% highlighting a notable concern for individuals with glaucoma that could impact treatment adherence and overall self-management in glaucoma. Our finding is similar to a U.S. study by Wang et al, who used the same screening tool (PHQ-9) and reported a prevalence of 10.9% (36). A case-control study in Brazil using HADS reported prevalence of depression at 10.1% (14), and another U.S. study using the GDS reported a prevalence of mild to moderate depressive symptoms at 12.2%, with none scoring for severe depression (12).

However, our prevalence appears to be relatively low in comparison to other studies. For example, a systematic review reported a pooled prevalence of depression at approximately 25% among people with glaucoma (37). In an Australian study, using the GDS-15 revealed rates of 11.4% for mild, 20.9% for moderate, and 32.1% for severe depressive symptoms (17). In India, a hospital-based cross-sectional study measuring depression using PHQ-9, recruited outpatients with glaucoma aged 40 – 80 years, the prevalence is as high as 35.8%, with 20.95% experiencing mild, 8.11% moderate, and 6.76% severe depressive symptoms (5). Other studies from China (15), Singapore (3), Turkey (11) also noted higher prevalence. Variations could be due to differences in the methodological approaches, tools used, and sample sizes.

In studies from Sub-Saharan Africa, particularly from Nigeria, where rates have ranged from 41.8% to 21.7% (13,18,19,38). These discrepancies may be attributed to methodological approaches, such as the use of a case-control design by Ubochi at al. (18) in contrast to our cross-sectional design. Variations in the age of participants may also play a role, as these previous studies focused primarily on older adults, whereas our study included a younger population as well. Additionally, cross-country variations in cultural beliefs, such as stigma, reliance on traditional healers or religious leaders rather than professional mental health services, and limited mental health awareness may contribute to the differing prevalence rates. Stigma around mental health, in particular, can discourage individuals from acknowledging or reporting depressive symptoms, leading to potential underestimation.

However, our results obtained higher rates than that reported by Rezapour et al in Germany, where the same tool (PHQ-9) was used at a cutoff point of ≥10 revealing overall prevalence of depression of 6.6% among participants with glaucoma with 24.3% having minimal, 4.9% moderate, 1.7% moderately severe, and none (0.0%) scored severe depressive symptoms (16). The discrepancy in prevalence may stem from differences in sample size and study population, as our study was a small, hospital-based sample, whereas theirs was a larger, population-based cohort. Our population might also have higher rates of depression compared to a setting from a high-income country due to cultural differences in how depression is understood and expressed despite use of validated tools, economic conditions of participants, and access to mental health services.

This study identified that participants with good social support were less likely to experience depression than those with poor social support; having moderate and strong social support appeared to be protective against depression. This finding aligns with a study by Yimer et al, which reported that individuals with visual impairments and inadequate social networks are at a higher risk for depression (25). Given that glaucoma is a progressive disease that gradually affects vision and limits daily functioning, social support becomes essential in buffering psychological stress associated with the disease. Social support can come from family, friends, or community members and may involve emotional, financial, and or practical assistance, which helps reduce stress and improve mental well-being.

Status of living arrangement was not a factor associated with probable depression unlike the finding from a Taiwanese study by Chen et al (24) where they found that living alone was a risk factor for developing depression. It is important to consider that the quality of the relationships with family members is equally important. Some individuals may live with family yet experience loneliness due to a lack of social engagement and interaction, which may potentially contribute to depressive symptoms (39,40). Conversely, individuals who live alone but maintain strong external connections may experience lower depressive symptoms. Therefore, both the presence and quality of social relationships are critical when assessing depression in people with glaucoma (40).

Although previous studies (5,6,15) revealed that elderly individuals with glaucoma had a higher likelihood of experiencing depression than the young, we observed no significant difference. The probable explanation could be due to the tool used (PHQ-9), which might not have picked up significant depressive symptoms among the elderly who mostly present with mood or cognitive symptoms that could have been picked up by a tool like GDS. It is also observed that older persons are less likely to report depressive symptoms due to either generational stigma in communities (41), medical conditions masking depression, or due to different symptom presentations (42).

Furthermore, in this Tanzanian sample, similar to findings from studies conducted in India, China, and Nigeria (5,15,19), the prevalence of probable depression did not significantly differ by sex. Although males had higher levels of probable depression than females in our study, the difference was not statistically significant, as also reported by Ajith et al.; the difference seen varied by chance (5). In contrast, other studies from Singapore, Nigeria, and Brazil (6,13,14,23), found significant sex-based differences, with females reporting more depressive symptoms than males. The discrepancy observed between studies could have stemmed from differences in the level of social support across study populations, as participants in our sample had better social support for both men and women and had equal access to care, which might have removed the sex-related difference in depressive symptoms.

In this study, the severity of glaucoma, assessed using an objective measure Vertical-Cup-to-Disc-Ratio (VCDR), was not significantly associated with probable depression. This result aligns with studies from the USA and China, where no association was found between clinically defined objective measures of visual function Best corrected Visual Acuity (BCVA), Mean Deviation (MD), Intra-Ocular Pressure (IOP), and Vertical-Cup-to-Disc-Ratio (VCDR) and depression, however they reported an association when subjective measure of visual function National Eye Institute of Visual Functioning Quality-25 (NEI-VFQ25) was used (15,36). In contrast, other studies from Australia, Japan, Singapore, and Nigeria using only objective measures, reported a significant association between glaucoma severity and depression (13,17,22,23). These discrepancies suggest that the choice of measurement tools for glaucoma severity may influence the observed relationship between glaucoma and depression. Objective clinical measures capture physiological severity of glaucoma but may not reflect the patient’s lived experience of visual impairment. Subjective assessments on the other hand, such as NEI-VFQ-25 consider how individuals perceive the impact of visual loss and their quality of life which can affect psychological well-being. For instance, Rezapour et al. emphasized that perceived visual disability, rather than clinical severity measures, may better predict depressive symptoms (16). Similarly, Diniz-Filho et al. found that rapid visual field loss increased the risk of depression and that changes in depressive symptoms were better predicted by assessing visual field loss than by disease severity alone (43).

Additionally, this study found no significant association between level of education or employment status with probable depression. These findings are consistent with previous research From Brazil, Singapore, and Nigeria (14,18,23,43). However, other studies have reported significant associations in education (15,19) and employment (13,19). These conflicting results may be explained by differences in study populations, sample sizes or access to health care bringing about the inconsistencies in the observed associations. In our study population, having stronger social support might have alleviated the impact of the psychological burden of unemployment or low educational attainment.

## Limitations

The study has several limitations that should be considered when interpreting the findings. First, majority of the participants were older and, therefore, they might have difficulty providing accurate information as questions require recalling information in a period past two weeks, which may be subject to recall bias. Although efforts were made to mitigate by interviewing in a private, comfortable room at a slow pace to reduce anxiety, improve recall, and ensure confidentiality. Secondly, the population studied in this research was rare during the specified period and was drawn from a single tertiary hospital in an urban setting. This selection of population from urban and tertiary hospitals might not reflect patients from the rest of Tanzania, where findings might be different in rural care. This might have introduced selection bias. Additionally, a consecutive sampling method was employed. While the findings of this study are generalizable within the specific context of MNH, their applicability to the broader community is limited.

Participants may have chosen to reveal information that seemed more socially acceptable due to feelings of shame, embarrassment, or fear of stigma relating to having depressive symptoms, and the level of social support introduces social desirability bias. This might affect the estimation of the prevalence. This was mitigated by well-educating the participants on the purpose of the interview and the benefits of the study, as well as ensuring anonymity and confidentiality of their participation, hence encouraging them to open up.

### 6.1 Conclusion and Recommendations

Depressive symptoms are an important concern among adults with glaucoma, with social support identified as a significant protective factor. This underscores the importance of regular mental health screenings in ophthalmology clinics using screening tools to identify individuals experiencing depression, who may benefit from psychological care. We recommend a multidisciplinary approach whereby ophthalmologists and primary health care workers integrate psychological services and screening of depression among those with glaucoma for early detection. Early identification is crucial for timely referrals and management of depression comorbidity, which can significantly impact a person’s quality of life and improve their treatment adherence. The progressive nature of this condition can lead to distressing consequences related to potential vision loss; thus, comprehensive care should address both physical and mental health needs.

Additionally, evidence from this study indicates that those who experience robust social support exhibit significantly lower levels of depressive symptoms compared to individuals with limited support networks. It is essential for our community to recognize the importance of providing both financial and emotional support to individuals with ocular diseases. Therefore, we advocate public health stakeholders to promote importance of social support for people with ocular diseases like glaucoma in communities through educational initiatives. We advocate expanding research across diverse settings, utilizing larger sample sizes, considering using tools specific for elderly like GDS as it was observed majority of those with glaucoma were elderly.

## Data Availability

The datasets generated and/or analyzed during the current study are not publicly available due to ongoing planned secondary analyses by the study investigators aimed at strengthening research capacity among collaborators based in a low- and middle-income country (Tanzania). The data are however, available from the corresponding author on reasonable request for researchers who meet the criteria for access to confidential data.

## Acknowledgements

We would like to extend our sincere thanks to all the participants who generously took part in the study. Their contributions have been invaluable in advancing knowledge in this field. Our sincere gratitude also goes to the research assistants for their hard work and commitment in collecting data. We appreciate Dr. Sayyada Shabbir Sachedina, who assisted in the methodological processes and queries relating to glaucoma. Lastly, we extend our gratitude to the administrative offices of the Department of Psychiatry and Mental Health staff (MUHAS and MNH) and the Department of Ophthalmology (staff and residents) for their exceptional assistance.

## Authors’ contribution

JSR developed the proposal and report. SM conducted the data analysis. SK and CM oversaw the development process and interpretation of results. ESM, ZM, and FEG guided data interpretation and manuscript writing. All authors reviewed and approved the final version of the manuscript.

## Supporting Information

**S1 Fig. Flow chart of participants through the study**

**S2 Fig. Prevalence of depressive symptoms among adults with glaucoma**

**S1 File. Consent Form. English version**

## References

1. Tham YC, Li X, Wong TY, Quigley HA, Aung T, Cheng CY. Global prevalence of glaucoma and projections of glaucoma burden through 2040: A systematic review and meta-analysis. Ophthalmology. 2014 Nov 1;121(11):2081–90. doi:10.1016/j.ophtha.2014.05.013 PubMed PMID: 24974815.

2. Study VLEG of the GB of D. Global estimates on the number of people blind or visually impaired by glaucoma: A meta-analysis from 2000 to 2020. Eye. 2024;38(11):2036.

3. Asiamah R, Kyei S, Owusu G, Agyiri PE. Prevalence of glaucoma in Africa: A systematic review and Bayesian meta-analysis. PLoS One. 2025;20(8):e0330567.

4. Buhrmann RR, Quigley HA, Barron Y, West SK, Oliva MS, Mmbaga BBO. Prevalence of glaucoma in a rural East African population. Invest Ophthalmol Vis Sci. 2000;41(1):40–8.

5. Ajith BS, Najeeb N, John A, Anima VN. Cross sectional study of depression, anxiety and quality of life in glaucoma patients at a tertiary centre in North Kerala. Indian J Ophthalmol. 2022;70(2):546.

6. Stamatiou ME, Kazantzis D, Theodossiadis P, Chatziralli I. Depression in glaucoma patients: A review of the literature. Semin Ophthalmol. 2022;37(1):29–35. doi:10.1080/08820538.2021.1903945 PubMed PMID: 33822676.

7. Chong J, Reinschmidt KM, Moreno FA. Symptoms of depression in a Hispanic primary care population with and without chronic medical illnesses. Prim Care Companion CNS Disord. 2010;12(3):26914.

8. Bedasso K, Bedaso A, Feyera F, Gebeyehu A, Yohannis Z. Prevalence of common mental disorders and associated factors among people with glaucoma attending outpatient clinic at Menelik II Referral Hospital, Addis Ababa, Ethiopia. PLoS One. 2016;11(9):e0161442.

9. Tilahun MM, Yibekal BT, Kerebih H, Ayele FA. Prevalence of common mental disorders and associated factors among adults with Glaucoma attending University of Gondar comprehensive specialized hospital tertiary eye care and training center, Northwest, Ethiopia 2020. PLoS One. 2021;16(5):e0252064.

10. Mabuchi F, Yoshimura K, Kashiwagi K, Shioe K, Yamagata Z, Kanba S, et al. High prevalence of anxiety and depression in patients with primary open-angle glaucoma. J Glaucoma. 2008;17(7):552–7.

11. Tastan S, Iyigun E, Bayer A, Acikel C. Anxiety, depression, and quality of life in Turkish patients with glaucoma. Psychol Rep. 2010;106(2):343–57.

12. Yochim BP, Mueller AE, Kane KD, Kahook MY. Prevalence of cognitive impairment, depression, and anxiety symptoms among older adults with glaucoma. J Glaucoma. 2012;21(4):250–4.

13. Onwubiko SN, Nwachukwu NZ, Muomah RC, Okoloagu NM, Ngwegu OM, Nwachukwu DC. Factors associated with depression and anxiety among glaucoma patients in a tertiary hospital South-East Nigeria. Niger J Clin Pract. 2020;23(3):315–21.

14. Abe RY, Silva LNP, Silva DM, Vasconcellos JPC, Costa VP. Prevalence of depressive and anxiety disorders in patients with glaucoma: a cross-sectional study. Arq Bras Oftalmol. 2021;84:31–6.

15. Wu N, Kong X, Sun X. Anxiety and depression in Chinese patients with glaucoma and its correlations with vision-related quality of life and visual function indices: a cross-sectional study. BMJ Open. 2022;12(2):e046194.

16. Rezapour J, Nickels S, Schuster AK, Michal M, Münzel T, Wild PS, et al. Prevalence of depression and anxiety among participants with glaucoma in a population-based cohort study: The Gutenberg Health Study. BMC Ophthalmol. 2018;18:1–9.

17. Skalicky S, Goldberg I. Depression and quality of life in patients with glaucoma: a cross-sectional analysis using the Geriatric Depression Scale-15, assessment of function related to vision, and the Glaucoma Quality of Life-15. J Glaucoma. 2008;17(7):546–51.

18. Ubochi CC, Achigbu EO, Nkwogu FU, Onyia OE, Okeke CJ. The impact of glaucoma on the mental health of primary open-angle glaucoma patients attending a teaching hospital in South East Nigeria. J West Afr Coll Surg. 2020;10(2):17.

19. Okudo AC, Babalola OE, Ogunro AT. A comparative analysis of anxiety and depression among glaucoma and cataractous patients in Southwest Nigeria. Open J Ophthalmol. 2021;11(02):105–33.

20. Greer TL, Kurian BT, Trivedi MH. Defining and Measuring Functional: Recovery from Depression. CNS Drugs. 2010;24:267–84.

21. Murdoch I, Smith AF, Baker H, Shilio B, Dhalla K. The cost and quality of life impact of glaucoma in Tanzania: An observational study. PLoS One. 2020;15(6):e0232796.

22. Mabuchi F, Yoshimura K, Kashiwagi K, Yamagata Z, Kanba S, Iijima H, et al. Risk factors for anxiety and depression in patients with glaucoma. British Journal of Ophthalmology. 2012;96(6):821–5.

23. Lim NCS, Fan CHJ, Yong MKH, Wong EPY, Yip LWY. Assessment of Depression, Anxiety, and Quality of Life in Singaporean Patients with Glaucoma. J Glaucoma. 2016 Jul 1;25(7):605–12. doi:10.1097/IJG.0000000000000393 PubMed PMID: 26950574.

24. Chen YY, Lai YJ, Wang JP, Shen YC, Wang CY, Chen HH, et al. The association between glaucoma and risk of depression: a nationwide population-based cohort study. BMC Ophthalmol. 2018;18:1–8.

25. Yimer YM, Buli MB, Nenko G, Mirkena Y, Kassew T. The prevalence and determinant factors of self-reported depressive symptoms among elderly people with visual impairment attending an outpatient clinic in Ethiopia. Clin Optom (Auckl). 2021;63–72.

26. Lundmark PO, Trope GE, Shapiro CM, Flanagan JG. Depressive symptomatology in tertiary-care glaucoma patients. Canadian Journal of Ophthalmology. 2009;44(2):198–204.

27. Ministry of Finance and Planning NB of S and POF and PO of the CGSZanzibar. The 2022 Population and Housing Census: Initial Results. Dodoma, Tanzania; 2022 Oct. Report.

28. Cochran WG. Sampling techniques. john wiley & sons; 1977.

29. Jung Y, Han K, Wang S min, Yoon H yeon, Moon J Il. Effect of depressive symptom and depressive disorder on glaucoma incidence in elderly. Sci Rep. 2021;11(1):5888.

30. Committee COSGPGE. Corrigendum: Canadian Ophthalmological Society evidence-based clinical practice guidelines for the management of glaucoma in the adult eye/Guide factuel de pratique clinique de la Société canadienne d’ophtalmologie pour la gestion du glaucome chez l’adulte. Canadian Journal of Ophthalmology. 2009;44(4):477.

31. Kroenke K, Spitzer RL, Williams JBW. The PHQ-9: validity of a brief depression severity measure. J Gen Intern Med. 2001;16(9):606–13.

32. Fawzi MCS, Ngakongwa F, Liu Y, Rutayuga T, Siril H, Somba M, et al. Validating the Patient Health Questionnaire-9 (PHQ-9) for screening of depression in Tanzania. Neurol Psychiatry Brain Res. 2019;31:9–14.

33. Kocalevent RD, Berg L, Beutel ME, Hinz A, Zenger M, Härter M, et al. Social support in the general population: standardization of the Oslo social support scale (OSSS-3). BMC Psychol. 2018;6(1):1–8.

34. Adams DJ, Ndanzi T, Rweyunga AP, George J, Mhando L, Ngocho JS, et al. Depression and associated factors among geriatric population in Moshi district council, Northern Tanzania. Aging Ment Health. 2021;25(6):1035–41.

35. Press S. Stata statistical software: Release 16. StataCorp LLC. 2019.

36. Wang SY, Singh K, Lin SC. Prevalence and predictors of depression among participants with glaucoma in a nationally representative population sample. Am J Ophthalmol. 2012;154(3):436–44.

37. Zheng Y, Wu X, Lin X, Lin H. The Prevalence of Depression and Depressive Symptoms among Eye Disease Patients: A Systematic Review and Meta-analysis. Scientific Reports. Nature Publishing Group; 2017. doi:10.1038/srep46453 PubMed PMID: 28401923.

38. Akindipe TO, Aina F, Onakoya AO. Risk of depression and subjective quality of life among attendees of a West African glaucoma clinic. Int J Med Med Sci. 2011;1(2):432–5.

39. Padayachey U, Ramlall S, Chipps J. Depression in older adults: prevalence and risk factors in a primary health care sample. South African family practice. 2017;59(2):61–6.

40. Ojagbemi A, Gureje O. Social relationships and the association of loneliness with major depressive disorder in the Ibadan study of aging. World Social Psychiatry. 2019;1(1):82–8.

41. Conner KO, Copeland VC, Grote NK, Rosen D, Albert S, McMurray ML, et al. Barriers to treatment and culturally endorsed coping strategies among depressed African-American older adults. Aging Ment Health. 2010;14(8):971–83.

42. Antony A, Parida SP, Behera P, Padhy SK. Geriatric depression: prevalence and its associated factors in rural Odisha. Front Public Health. 2023;11:1180446.

43. Diniz-Filho A, Abe RY, Cho HJ, Baig S, Gracitelli CPB, Medeiros FA. Fast visual field progression is associated with depressive symptoms in patients with glaucoma. Ophthalmology. 2016;123(4):754–9.

